# Potential impacts of prolonged absence of influenza virus circulation on subsequent epidemics

**DOI:** 10.1101/2022.02.05.22270494

**Authors:** Simon P. J. de Jong, Zandra C. Felix Garza, Joseph C. Gibson, Alvin X. Han, Sarah van Leeuwen, Robert P. de Vries, Geert-Jan Boons, Marliek van Hoesel, Karen de Haan, Laura E. van Groeningen, Katina D. Hulme, Hugo D. G. van Willigen, Elke Wynberg, Godelieve J. de Bree, Amy Matser, Margreet Bakker, Lia van der Hoek, Maria Prins, Neeltje A. Kootstra, Dirk Eggink, Brooke E. Nichols, Menno D. de Jong, Colin A. Russell

## Abstract

**Background:** During the first two years of the COVID-19 pandemic, the circulation of seasonal influenza viruses was unprecedentedly low. This led to concerns that the lack of immune stimulation to influenza viruses combined with waning antibody titres could lead to increased susceptibility to influenza in subsequent seasons, resulting in larger and more severe epidemics.

**Methods:** We analyzed historical influenza virus epidemiological data from 2003-2019 to assess the historical frequency of near-absence of seasonal influenza virus circulation and its impact on the size and severity of subsequent epidemics. Additionally, we measured haemagglutination inhibition-based antibody titres against seasonal influenza viruses using longitudinal serum samples from 165 healthy adults, collected before and during the COVID-19 pandemic, and estimated how antibody titres against seasonal influenza waned during the first two years of the pandemic.

**Findings:** Low country-level prevalence of influenza virus (sub)types over one or more years occurred frequently before the COVID-19 pandemic and had relatively small impacts on subsequent epidemic size and severity. Additionally, antibody titres against seasonal influenza viruses waned negligibly during the first two years of the pandemic.

**Interpretation:** The commonly held notion that lulls in influenza virus circulation, as observed during the COVID-19 pandemic, will lead to larger and/or more severe subsequent epidemics might not be fully warranted, and it is likely that post-lull seasons will be similar in size and severity to pre-lull seasons.

**Funding:** European Research Council, Netherlands Organization for Scientific Research, Royal Dutch Academy of Sciences, Public Health Service of Amsterdam.

**Research in context:** *Evidence before this study:* During the first years of the COVID-19 pandemic, the incidence of seasonal influenza was unusually low, leading to widespread concerns of exceptionally large and/or severe influenza epidemics in the coming years. We searched PubMed and Google Scholar using a combination of search terms (i.e., “seasonal influenza”, “SARS-CoV-2”, “COVID-19”, “low incidence”, “waning rates”, “immune protection”) and critically considered published articles and preprints that studied or reviewed the low incidence of seasonal influenza viruses since the start of the COVID-19 pandemic and its potential impact on future seasonal influenza epidemics. We found a substantial body of work describing how influenza virus circulation was reduced during the COVID-19 pandemic, and a number of studies projecting the size of future epidemics, each positing that post-pandemic epidemics are likely to be larger than those observed pre-pandemic. However, it remains unclear to what extent the assumed relationship between accumulated susceptibility and subsequent epidemic size holds, and it remains unknown to what extent antibody levels have waned during the COVID-19 pandemic. Both are potentially crucial for accurate prediction of post-pandemic epidemic sizes.

*Added value of this study:* We find that the relationship between epidemic size and severity and the magnitude of circulation in the preceding season(s) is decidedly more complex than assumed, with the magnitude of influenza circulation in preceding seasons having only limited effects on subsequent epidemic size and severity. Rather, epidemic size and severity are dominated by season-specific effects unrelated to the magnitude of circulation in the preceding season(s). Similarly, we find that antibody levels waned only modestly during the COVID-19 pandemic.

*Implications of all the available evidence:* The lack of changes observed in the patterns of measured antibody titres against seasonal influenza viruses in adults and nearly two decades of epidemiological data suggest that post-pandemic epidemic sizes will likely be similar to those observed pre-pandemic, and challenge the commonly held notion that the widespread concern that the near-absence of seasonal influenza virus circulation during the COVID-19 pandemic, or potential future lulls, are likely to result in larger influenza epidemics in subsequent years.

## Introduction

Seasonal influenza viruses typically cause annual epidemics worldwide, infecting up to 35% of the human population.^1,2^ However, the incidence of seasonal influenza was unusually low during the first two years of the COVID-19 pandemic,^3,4^ likely due to non-pharmaceutical interventions (NPIs) aimed at reducing transmission and spread of SARS-CoV-2, which are also effective in limiting exposure to seasonal influenza viruses.^5–8^ This global lull in influenza virus circulation and consequent lack of immune stimulation led to widespread concerns of increased susceptibility to seasonal influenza viruses due to waning immunity, potentially resulting in larger and more severe epidemics in upcoming seasons, with studies predicting substantial increases in epidemic size.^6,9–12^ Importantly, however, these studies fundamentally rely on assumptions concerning the relationship between accumulated susceptibility and influenza epidemic size. Similarly, the results of these studies are strongly influenced by assumed, but unmeasured, dynamics of antibody waning within such a period of abnormally reduced circulation.

Here, we performed a data-driven investigation into the extent to which prolonged periods of absence of influenza virus circulation, such as seen during the COVID-19 pandemic, can be expected to lead to larger and more severe subsequent epidemics. First, we analyzed two decades of epidemiological data from 47 countries to investigate the frequency of influenza lulls in the past and their effects on subsequent epidemic size and severity. Second, to interrogate the effects of lulls in influenza virus circulation on influenza antibody dynamics, we investigated the extent to which antibody titres against seasonal influenza viruses waned during the COVID-19 pandemic, using serum samples collected longitudinally before and during the COVID-19 pandemic from adults living in the Netherlands.

## Methods

### Epidemiological analysis

To gain insights into the expected effects of influenza circulation lulls on post-lull influenza epidemic sizes, we investigated the effects that past (sub)type lulls had on subsequent (sub)type epidemic size. We analyzed virological surveillance data for 47 countries in the Northern and Southern Hemispheres for the period from 2002 until 2019, deposited in the WHO FluNet database.^13^ We identified (sub)type lull periods as consecutive seasons in which a particular (sub)type was not dominant, for a particular country, where we defined (sub)type dominance as a (sub)type accounting for ≥30% of detections in a particular country, in a particular season. Here, a (sub)type being dominant in two consecutive seasons corresponds to a lull duration of one.

Additionally, we estimated (sub)type-specific relative epidemic sizes for 20 countries in Europe and the Middle East by integrating virological surveillance data with influenza-like illness (ILI) data from the WHO FluID database.^14^ In these estimates, a relative size of one corresponds to the mean number of influenza virus infections in a single season for a given country, irrespective of (sub)type.

We used Bayesian hierarchical models to investigate the effects of multiple metrics of the magnitude of influenza virus circulation in preceding years on relative epidemic size. To investigate the effect of magnitude of prior incidence on epidemic severity, we compared our computed lull durations to estimated rates of Europe-wide influenza-specific excess mortality as calculated prior by the EuroMOMO network,^15,16^ which we use as a proxy for severity. Please see the Supplementary Appendix for full methodological details of all analyses outlined here.

### Antibody model

To investigate antibody dynamics during the COVID-19 pandemic, we measured antibody titres with haemagglutination inhibition (HI) assay against representative strains of each (sub)type of seasonal influenza (A/H3N2: A/Netherlands/04189/2017; A/H1N1pdm09: A/Netherlands/10218/2018; B/Yamagata: B/Netherlands/04136/2017; B/Victoria: B/Netherlands/00302/2018), in longitudinal serum samples collected in the summer of 2020 and 2021 from 100 male adults within the Amsterdam Cohort Studies on HIV infection and AIDS (ACS),^17^ as well as 130 serum samples from a longitudinal cohort of adult COVID-19 patients (the Viro-immunological, clinical and psychosocial correlates of disease severity and long-term outcomes of infection in SARS-CoV-2 – a prospective cohort study (RECoVERED)).^18^ To compare intra-pandemic against pre-pandemic influenza antibody dynamics, we additionally measured HI titres to the same strains for the same ACS individuals using serum samples collected in the summer of 2017, 2018 and 2019, yielding a total of 630 serum samples across both cohorts. Importantly, all ACS individuals were HIV-seronegative. Individuals from the RECoVERED cohort were confirmed to have not been vaccinated for influenza in 2020.

We used a mathematical model to estimate antibody waning rates, based on the measured haemagglutination inhibition antibody titres, with a Markov Chain Monte Carlo (MCMC) algorithm used to explore the distribution of model parameters and augmented data. Full details on virus selection, virus propagation, cohort details, HI assay used and the antibody waning model can be found in the Supplementary Appendix.

## Results

### Effects of past (sub)type lulls on subsequent (sub)type epidemic size

Prior to the COVID-19 pandemic, seasonal influenza virus circulation was highly heterogeneous, with individual influenza epidemics in any given country typically being dominated by one or two influenza virus (sub)types, leading to frequent lull periods lasting 1-3 years where other seasonal influenza virus subtypes barely circulated. Due to the lack of immunological cross-reactivity between (sub)types, these lulls are potentially analogous to the scenario observed in the first two years of the COVID-19 pandemic for individual (sub)types. Hence, they should yield insight into the effects of these lulls on epidemic size.

Between 2003 and 2020, low or near-absent circulation of an influenza virus (sub)type within a single season occurred frequently, accounting for 41%, 48%, and 62% of all country-season pairs for A/H3N2, A/H1N1pdm09 and influenza B viruses, respectively. In 53%, 56%, and 88% of country-season pairs for A/H3N2, A/H1N1pdm09, and B viruses, respectively, (sub)type lulls lasted for more than one year, with some lull periods lasting as long as three years (Fig. 1a). This indicates that extended periods of relative absence of individual influenza (sub)types are a regular feature of influenza epidemic dynamics. Very small or absent (sub)type-specific epidemics (*i*.*e*. relative epidemic sizes < 0·1, corresponding to 10% of the mean influenza epidemic size) were observed for 28%, 23%, and 37% of country-seasons for A/H3N2, A/H1N1pdm09 and influenza B viruses, respectively (Fig. 1b).

**Fig. 1.**
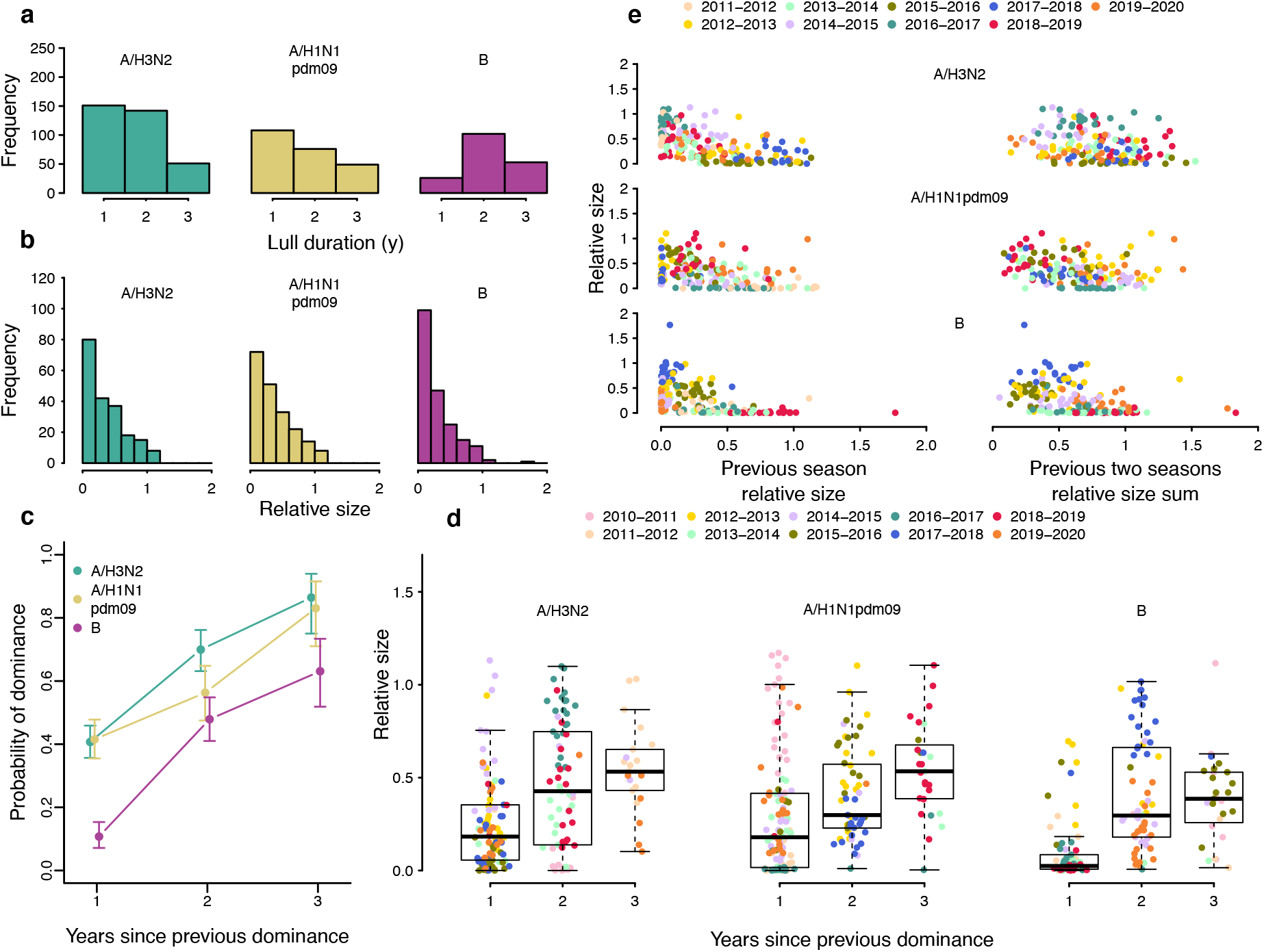
The effects of previous years’ influenza virus circulation on epidemic size and composition. **a**, The distribution of lull period durations, by (sub)type, across all country-season pairs. A lull duration of one corresponds to the same (sub)type’s dominance in the previous season and presence in the current season. **b**, The distribution of relative epidemic sizes by virus (sub)type, across all countries and seasons. **c**, The probability of (sub)type dominance as a function of years since previous dominance. Error bars correspond to 95% confidence interval from an exact two-tailed binomial test for proportions. **d**, Relative size of a (sub)type-specific epidemic as a function of number of years since previous dominance of that (sub)type in that country, colored by season. Each point corresponds to a country-season pair, colored by the season. **e**, Relative size of a subtype’s epidemic as a function of its size in the previous season and the sum of the two previous seasons’ sizes. Each point corresponds to a country-season pair, colored by the season.

Investigating the effect influenza virus (sub)type lulls had on epidemic composition and size, we found that although both the probability of a (sub)type’s dominance and the mean epidemic sizes for each influenza virus (sub)type increased with time since previous dominance (Fig. 1c-d), epidemic sizes varied substantially for each value of years since last dominance (Fig. 1d). This suggests that background variation in epidemic size, independent of absence or presence of circulation in preceding years, is substantial. Similarly, while there is a negative relationship between the relative epidemic size of each (sub)type to its relative size in the preceding year and the relative summed size over the last two years, there is wide variation in epidemic size: seasons with very low and very high relative sizes both occurred frequently following years of low-to-mid incidence (Fig. 1e). Notably, in 9 of the 20 countries included in our dataset, the first A/H3N2-dominant season (2011/2012) after the 2009 A/H1N1pdm09 pandemic did not belong to the three largest A/H3N2 epidemics in the influenza seasons from 2010/2011 until 2019/2020, despite three years of near-absent circulation.

Importantly, for each number of years since dominance, we observed a striking degree of clustering of relative epidemic sizes across countries by season, suggesting the existence of season-specific effects on epidemic size, shared among countries in a single season (Fig. 1d). For example, in the 2013/2014 and 2016/2017 seasons, where A/H3N2 dominated in most countries two years prior, the relative incidence in 2016/2017 appeared consistently higher than in 2013/2014. We thus hypothesized that the size of (sub)type-specific epidemics could be jointly explained by a combination of season-specific effects shared among countries and effects related to the presence or absence of that virus (sub)type in the years preceding an epidemic. We used a Bayesian hierarchical model to estimate the likely effects of years since dominance, size in the previous year and the sum of previous two seasons’ sizes, as well as season-specific effects, on epidemic size (Fig. 2). The season effects correspond to the predicted ‘base size’ of a country’s epidemic in a particular season, independent of the magnitude of prior circulation. Each of the three predictors individually had non-trivial effects on epidemic size in models with season effects and estimated effects were substantially smaller than in models that did not include season effects (Fig. 2).

**Fig. 2.**
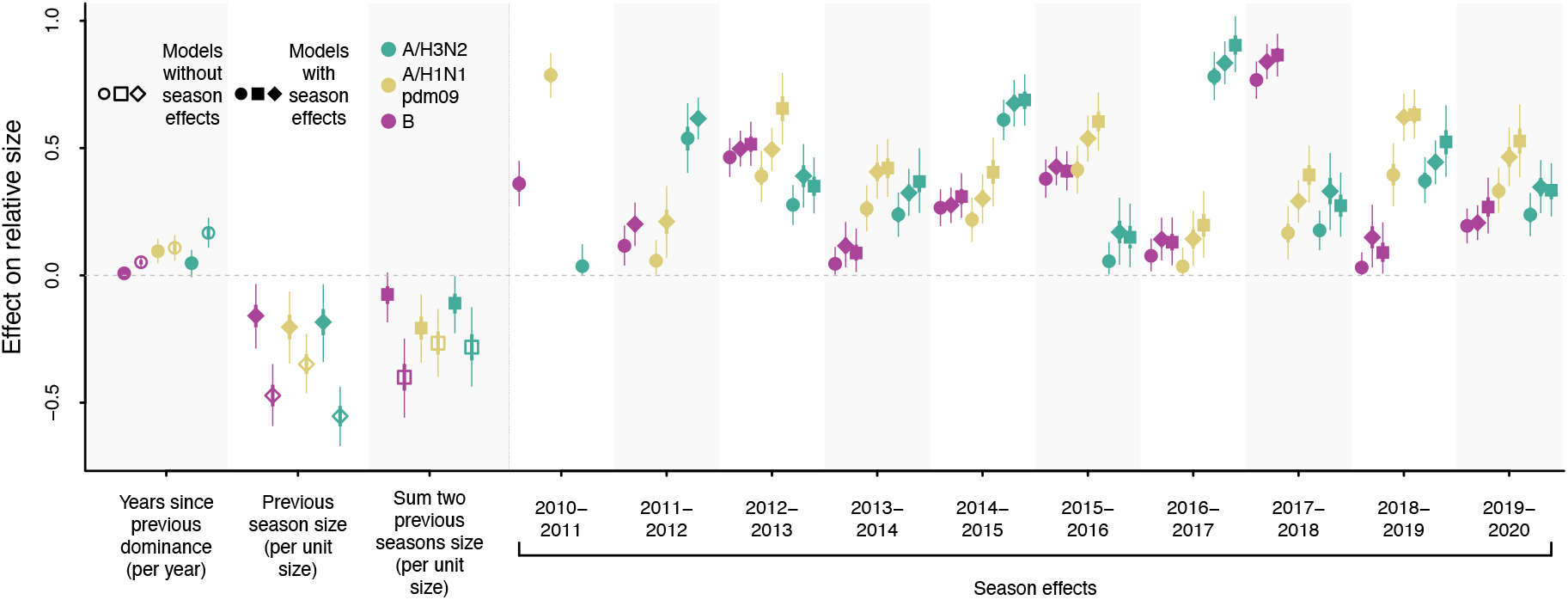
Bayesian hierarchical model correlating relative influenza (sub)type epidemic size to time since previous dominance, previous epidemic sizes and season-specific effects. Posterior distributions of parameter estimates in the model, with one year since previous dominance (circles), previous epidemic size (diamonds), or sum of previous two epidemics’ size (squares) as predictors, either with or without season effects. Points, thick and thin lines correspond to the posterior mean, 50% CI, and 95% CIs, respectively.

Crucially, across all model formulations, the estimated season effects, shared among countries, differed substantially between seasons. Models that included season effects exhibited much better predictive performance than models without season effects (Supplementary Fig. 1), and between-season differences with regard to season effects were consistently substantially greater in magnitude than any of the predictors related to prior incidence. For example, in the model that includes previous season size as predictor for A/H3N2 epidemic size, the estimated season effects (‘base sizes’) ranged from 0·17 (95% CI 0·04-0·31) in 2015/2016 to 0·83 (95% CI 0·75-0·92) in 2016/2017: a difference of 0·66.

Conversely, assuming the size of the previous season was the mean A/H3N2 season relative size (across all included countries and seasons), the effect of previous season size would only decrease predicted size by 0·06 (95% CI 0·01-0·12) compared to if there were no circulation in the previous season. Together, these results suggest that there is only a limited impact of the magnitude of influenza virus circulation in the preceding season(s) on subsequent epidemic size, consistent with previous work,^19^ and that epidemic size is dominated by season-specific factors, unrelated to the magnitude of prior circulation.

### Effects of past (sub)type lulls on subsequent influenza season severity

To investigate the effect of low influenza virus circulation on subsequent influenza season severity, as opposed to size, we compared our computed lull durations to Europe-wide estimates of excess mortality as calculated by the EuroMOMO network.^15,16^ Rates of pooled Europe-wide influenza-attribute excess mortality varied substantially between seasons, ranging from 0·31 (95% CI 0·24-0·38) per 100,000 in 2013/2014 to 28·58 (95% CI: 28·22-28·95) per 100,000 in 2014/2015. Hence, in the decade prior to the COVID-19 pandemic, epidemics could differ by up to two orders of magnitude in their severity.^15,16^ In the 2011/2012 season, which was A/H3N2-dominant Europe-wide and followed a three-year A/H3N2 lull in almost all countries, Europe-wide total excess mortality in the winter period amounted to 6·73 (95% CI 5·26-8·21) per 100,000. In turn, in the 2014/2015 and 2016/2017 seasons, which were also A/H3N2-dominant Europe-wide and followed lull periods of one and two years in almost all countries, respectively, influenza-specific excess mortality amounted to 28·58 (95% CI: 28·22-28·95) and 25·65 (95% CI: 25·26-26·05) per 100,000, respectively.^15,16^ Hence, in these seasons, influenza-specific excess mortality was four-to-five fold higher than total winter period excess mortality in 2011/2012, despite substantially shorter lull durations. While this coarse analysis can only be performed for seasons dominated by a single (sub)type, these results suggest that there is no clear relationship between the magnitude of circulation in the preceding seasons and the severity of subsequent seasons.

### Antibody responses to seasonal influenza virus during the COVID-19 pandemic

Waning of pre-existing immunity due to lack of immune stimulation has been posited to lead to larger post-lull epidemics, but evidence is lacking on precisely to what degree antibody immunity against seasonal influenza viruses might change during near-absence of seasonal influenza, as seen in the COVID-19 pandemic. To explicitly quantify the effects of lack of influenza virus circulation on antibody titres against seasonal influenza viruses, we performed an analysis of influenza antibody dynamics in the pre- and intra-COVID-19 pandemic period in the Netherlands. We quantified the baseline antibody titres of an adult population in the Netherlands for the seasons preceding the COVID-19 pandemic and the extent of their decrease during the pandemic. Importantly, antibody responses to the haemagglutinin protein of influenza viruses are known to be correlates of protection.^20–22^

From 2019 to 2021, mean HI titres remained largely unchanged for all influenza virus (sub)types, including during the COVID-19 pandemic period, for both the ACS and RECoVERED cohorts (Fig. 3b, Supplementary Fig. 4b). Influenza A/H3N2, A/H1N1pdm09 and B/Yamagata viruses had caused epidemics in the three influenza seasons prior to the onset of the COVID-19 pandemic (Fig. 3a) and epidemic activity during this period was consistent with patterns from 2010-2019 (Supplementary Fig. 2). For all seasonal influenza virus (sub)types, mean HI titres increased after the 2017/2018 influenza epidemic but returned to pre-2017/2018 levels by summer 2019 in the ACS cohort (Fig. 3b, Supplementary Fig. 4b). Differentiating the year-on-year individual HI titre distributions by titre rises that are indicative of recent influenza virus infection (≥4-fold increase, ≥2 log_2_ units), showed that influenza A and B virus infections were most common in individuals with low antibody titres in the year prior to infection (Fig. 3c, Supplementary Fig. 4c); consistent with lower antibody titers being associated with greater risk of infection. Overall, the HI titre distributions of the cohort remained largely unchanged over the study period, including during the first two years of the COVID-19 pandemic.

**Fig. 3.**
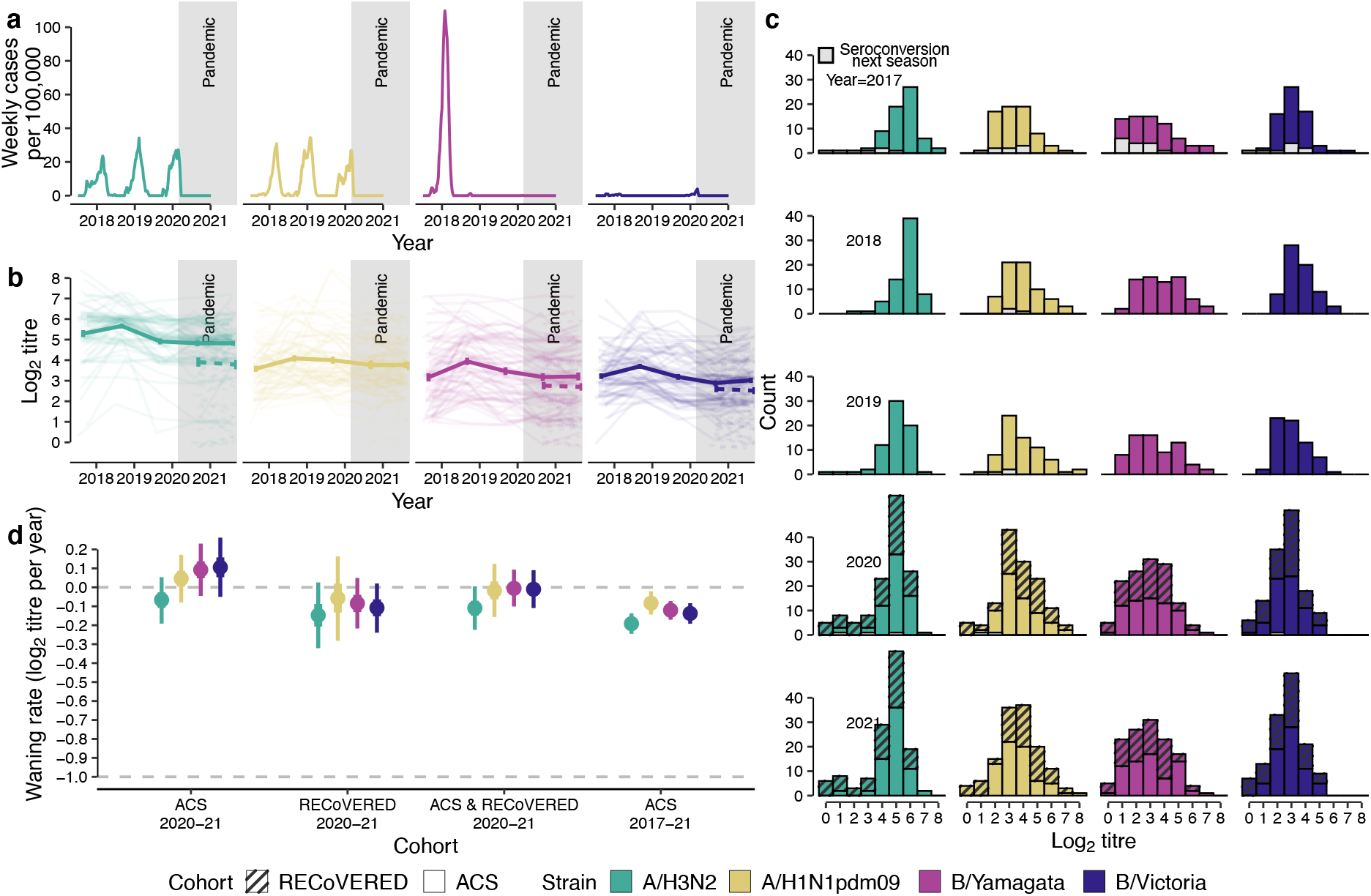
Waning antibody titres to seasonal influenza virus before and during the COVID-19 pandemic. **a**, Individual antibody titres against seasonal influenza viruses based on haemagglutination inhibition (HI) assay from 2017-2021 among 70 healthy male adult participants of the Amsterdam Cohort Studies on HIV infection and AIDS (ACS) cohort for each influenza virus (sub)type as well as 65 male and female participants of the RECoVERED cohort for years 2020-21 (dashed). Mean antibody tires changes across all individuals are drawn in bold lines with error bars indicating the mean standard error. **b**, Seasonal influenza virus epidemic activity 2017-2021 in the Netherlands based on virological and syndromic surveillance data. **c**, HI titre distributions in the two cohorts following each winter epidemic period colored by influenza virus (sub)type. HI titre distributions of individuals who experienced a ≥2 log_2_ units increase in HI titre (≥4-fold increase in HI titre), indicating likely infection in the next winter epidemic period, are shown in grey bars. **d**, Mean HI antibody titre waning rates by influenza virus (sub)type in adults estimated from HI titres from 70 ACS and 65 RECoVERED participants. Error bars correspond to the 50% and 95% credible interval from the Markov Chain Monte Carlo algorithm used to explore the distribution of model parameters. Waning rate of -1.0 corresponds to one two-fold decrease in antibody titre in one year.

**Fig. 4.**
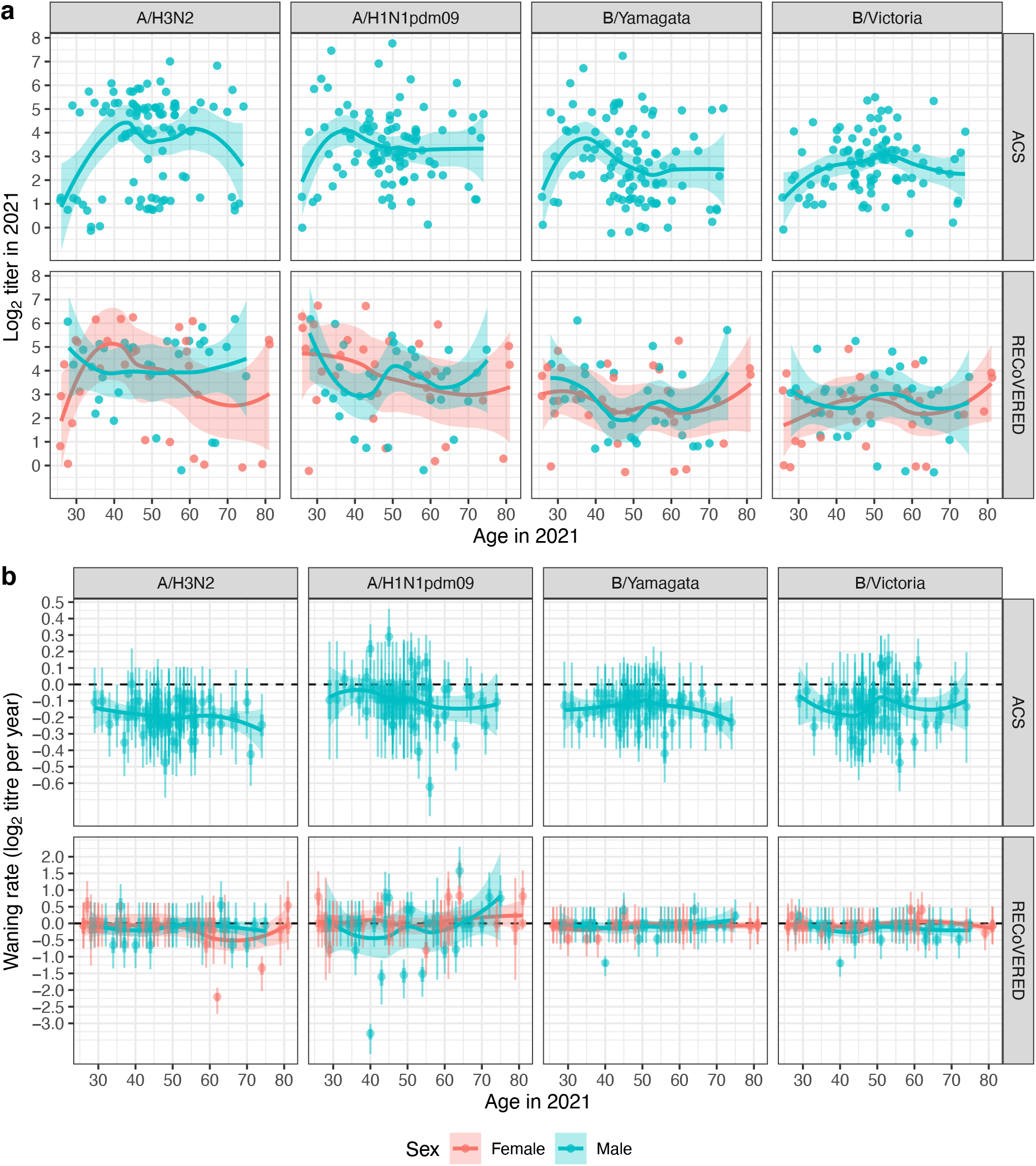
The effects of age and sex on baseline antibody titre and waning rate. **a**, A cross section of antibody titres for both cohorts in 2021, broken down by (sub)type, age and sex. **b**, Individual fitted waning rates with 50% (thick lines) and 95% (narrow lines) CIs for the ACS 2017-21 and RECoVERED 2020-21 data, broken down by strain, age and sex.

We applied a mathematical model on the HI titres of participants in 2020 and 2021 to estimate pandemic-period antibody titre waning rates. For the ACS individuals, we estimated that antibody titres against A/H3N2 viruses waned at -0·20 log_2_ units per year, 95% credible interval (CI) (−0·24, -0·16); A/H1N1pdm09 viruses at -0·10, 95% CI (−0·12, -0·07); B/Victoria viruses at -0·13, 95% CI (−0·16, -0·10); and B/Yamagata viruses at -0·14, 95% CI (−0·17, -0·11) (Fig. 3d, Supplementary Fig. 4d). For the RECoVERED cohort, we estimated mean waning rates towards A/H3N2, A/H1N1pdm09, B/Yamagata, and B/Victoria to be - 0·15, 95% CI (−0·31, 0·01), -0·08, 95% CI (−0·19, 0·03), -0·08, 95% CI (−0·20, 0·04) and - 0·10, 95% CI (−0·22, 0·02) log_2_ units per year respectively, in agreement with those derived from the ACS cohort (Fig. 3d). Combining data from both cohorts for the 2020-2021 period, the estimated mean waning rates remained similar to previous estimates for A/H3N2, A/H1N1pdm09 and B/Yamagata, and negligible for B/Victoria. We also calculated mean waning rates using HI titres from the same ACS individuals for the entire 2017-2021 period (Fig. 3d, Supplementary Fig. 4d), including only individuals who were likely not infected during the 2017-2021 period (*i*.*e*. no ≥2 log_2_ unit increases in HI titre for the entire study period). For this period, no significant waning of HI titres against any of the viruses was observed either, and estimates were similar to estimates for the 2020-2021 period, for both the ACS and the RECoVERED cohorts. We stratified baseline antibody titres by age and sex, for each (sub)type, but found no consistent age- or sex-related effects on baseline titres (Fig. 4a) or antibody waning rates (Fig. 4b). Measurement error was found to be consistent in both datasets at 0·38, 95% CI (0·36, 0·40) and 0·33, 95% CI (0·31, 0·36) log_2_ units for the full ACS and RECoVERED cohorts respectively, corresponding to a one-sided probability of a 2-fold error of approximately 5 −11%.

## Discussion

Our analysis of two decades of epidemiological data from 47 countries demonstrates that low country-level prevalence of influenza (sub)types over one or more years was not unique to the COVID-19 pandemic and occurred frequently in the past, and that periods or low or near-absent circulation of particular (sub)types did not necessarily lead to substantially increased epidemic sizes. Additionally, Bayesian statistical modelling shows that the magnitude of a (sub)type’s circulation in preceding years had only limited effect on subsequent size. Instead, the strong clustering of different countries’ epidemic size within particular seasons, supported by statistical modelling, suggests that epidemic size is more likely influenced by season-specific effects that are unrelated to the absence or presence of circulation in the prior season(s). The precise determinants of these season effects are likely manifold, including factors like the flux of viral seeding, heterosubtypic competition, and antigenic novelty.^19,23,24^ Similarly, the severity of influenza seasons appears to be largely independent of the magnitude of influenza virus circulation in the preceding seasons. Importantly, this means that even if there were substantial accumulation of susceptibility, for example during the COVID-19 pandemic, it is likely that its effect on epidemic size and severity would be dwarfed by inherent season-to-season variation in epidemic size, unrelated to the absence or presence of substantial circulation in preceding years.

We showed that HI-measured immune protection against recent seasonal influenza viruses remained largely unchanged in adults since the start of the COVID-19 pandemic. Our analysis also suggests that substantial waning of antibody titres against seasonal influenza viruses occurs at timescales substantially longer than the lull in seasonal influenza virus circulation during the first two years of the COVID-19 pandemic^10^. Crucially, this analysis demonstrates that waning rates previously used to project post-COVID-19 lull epidemic sizes (e.g. waning of immunity within a single year^9^, two to four years^6^, or forty weeks^12^) are too high, and that waning rather happens on longer timescales, even following periods of absent circulation, in agreement with waning rates previously reported for adults during regular periods of influenza virus circulation.^25^

The serum samples were collected in two independent cohorts, with substantial diversity in age and sex, accounting for the elderly but excluding children. Due to the complex effects of immunosenescence, the elderly potentially exhibit differing antibody dynamics. Whilst studies have shown that vaccine-mediated protection wanes modestly quicker in those over 65 years of age,^26,27^ there is little evidence to support the notion that serum antibodies wane significantly faster for this age group. Our results showed similar antibody baseline titres and waning rates for adults below and above 65 years of age, suggesting that serum antibodies in both subgroups wane at similar rates.

Due to the lack of children in our serological analysis, the extent to which their waning rates have changed since the start of the COVID-19 pandemic remains uncertain. Immune dynamics in children are known to differ from those in adults,^28^ with potentially higher waning rates, which could lead to increased susceptibility to infection. Furthermore, the accrual of additional birth cohorts during prolonged periods of absence of influenza virus circulation might affect epidemic dynamics. However, the same dynamics of waning in children and population turnover also occurred in pre-pandemic (sub)type lulls and are thus fully incorporated in our epidemiological analyses. As such, the absence of child sera is unlikely to bias our conclusions.

We used influenza-like illness data from the WHO FluID database and virologically confirmed data from the WHO FluNet database in our epidemiological analyses. Bias might affect both data sources. In particular, the FluID ILI data is non-influenza-specific, and FluNet data might be biased due to e.g. the presence of convenience samples and overrepresentation of outpatient surveillance. However, the observed consistency in the estimated (sub)type-specific epidemic sizes across the 20 countries included in the analysis for any given season suggests that these data sources broadly capture influenza epidemiological dynamics. Therefore, our results are unlikely to be substantially affected by potential year-on-year differences in reporting behavior or unrepresentative sampling. Additionally, the applicability of our analysis to the post-COVID-19 pandemic-like situation is predicated on the absence of substantial heterosubtypic immunity. Importantly, heterosubtypic protection has previously been estimated to be exceedingly short-lived, with duration on the order of a single day.^28^ While we could only perform our severity analysis using Europe-wide excess mortality data, the clustering of epidemic sizes within seasons across European countries as observed in our epidemiological analysis suggests that, for any given season, Europe-wide severity data is likely representative of the country level.

Although participants in the RECoVERED cohort were confirmed to be unvaccinated during the study period, vaccination status for the ACS cohort was not known. However, in the Netherlands individuals <60 years of age are only eligible for influenza vaccination if they have underlying health conditions, and assuming population-wide rates of influenza vaccine uptake in the Netherlands, only 3% of the ACS individuals, who importantly were all HIV-seronegative, would be expected to be vaccinated. This, combined with the similarity in antibody baseline titres and waning rates when comparing adults below and above 60 years of age, suggests that lack of vaccination status for the ACS cohort is unlikely to bias our conclusions.

Caution is required when looking at the size of post-pandemic seasonal influenza epidemics as the COVID-19 pandemic has brought about immense changes in testing behavior, which render the direct comparison of epidemic sizes before and immediately after the pandemic difficult. However, preliminary insight into the effects of COVID-19 pandemic related absence of circulation can be gained from Australia, where surveillance data shows that the 2022 influenza season was not greater in size than the range of epidemic sizes observed in the decade prior to the COVID-19 pandemic.^29^

Past studies into the possible effects of a lull in influenza circulation on subsequent epidemic size have assumed that the relationship between accumulation of susceptibility and epidemic size can be predicted using standard SIR-type epidemic models. Here, we show that this relationship is decidedly more complex, and that season-to-season variation in epidemic size is dominated by factors not captured in current SIR models, with only a relatively small effect of the magnitude of epidemics in the preceding year(s). Additionally, our results show that antibody titres to influenza viruses waned marginally during the COVID-19 pandemic, and to a smaller extent than assumed in modelling studies. Using multiple sources of data to add nuance to a complex issue, our results challenge the commonly held notion that post-lull influenza epidemics will be substantially larger and/or more severe.

## Data Availability

All data produced are available online at the project GitHub repository.

https://github.com/AMC-LAEB/waning-immunity-to-flu

## Contributors

S.P.d.J., Z.C.F.G., J.C.G., A.X.H., D.E., M.D.d.J., and C.A.R. designed the research; Z.C.F.G, S.v.L., M.v.H., K.d.H., and L.E.v.G executed the experimental work; Z.C.F.G. and S.v.L. generated the antibody titre data; E.W., G.J.d.B, H.D.G.v.W., A.M., M.B., L.v.d.H., M.P., N.K., and M.D.d.J. collected the clinical samples; R.P.d.V. and G.J.B. made and provided the glycan remodelled turkey red blood cells; S.P.d.J. and J.C.G. implemented the modelling work and performed the data analysis; S.P.d.J.,Z.C.F.G., J.C.G., A.X.H., K.D.H., B.E.N., and C.A.R. wrote the first draft of the paper. All authors contributed to the critical revision of the paper.

## Data sharing

All of the de-identified raw hemagglutination inhibition data, as well as accession codes for GISAID data, used in this paper is provided as supplementary information files. Raw surveillance data downloaded from WHO FluNet and FluID can be found in the project GitHub repository (https://github.com/AMC-LAEB/waning-immunity-to-flu). Biological materials are available for study via the Amsterdam Cohort Studies on HIV infection and AIDS (ACS) and the Viro-immunological, clinical and psychosocial correlates of disease severity and long-term outcomes of infection in SARS-CoV-2 – a prospective cohort study (RECoVERED). Custom scripts used for data analysis and modelling are available at the project GitHub repository (https://github.com/AMC-LAEB/waning-immunity-to-flu). Any additional information required to reanalyze the data reported in this paper is available from the lead contact upon request.

## Acknowledgements

A.X.H., Z.C.F.G. and C.A.R. were supported by ERC NaviFlu (No. 818353). J.G. and C.A.R. were supported by NIH R01 (5R01AI132362-04). C.A.R. was also supported by an NWO Vici Award (09150182010027). R.P.dV. was supported by ERC starting grant 802780 and a Beijerinck Premium of the Royal Dutch Academy of Sciences. GJB was supported by the Netherlands Organization for Scientific Research (NWO TOPPUNT 718.015.003) and by an ERC advanced grant (101020769). The RECoVERED cohort is supported by NWO ZonMw (No. 10150062010002) and the Public Health Service of Amsterdam (Research & Development grant number 21-14). The Amsterdam Cohort Studies on HIV infection and AIDS, a collaboration between the Public Health Service Amsterdam, the Amsterdam UMC of the University of Amsterdam, Medical Center Jan van Goyen and the HIV Focus Center of the DC-Clinics, are part of the Netherlands HIV Monitoring Foundation and financially supported by the Center for Infectious Disease Control of the Netherlands National Institute for Public Health and the Environment. We gratefully acknowledge the authors and originating and submitting laboratories (supplementary information) for the reference sequences retrieved from GISAID’s EpiFlu Database used in this study. The authors thank all ACS and RECoVERED study participants. We are also grateful to Mr. Reinier van der Palen of the department of Chemical Biology and Drug Discovery, Utrecht University for his practical assistance with turkey erythrocyte glycan remodeling.

## References

1 Petrova VN, Russell CA. The evolution of seasonal influenza viruses. Nat Rev Microbiol 2017; 16: 47–60.

2 Cohen C, Kleynhans J, Moyes J, et al. Asymptomatic transmission and high community burden of seasonal influenza in an urban and a rural community in South Africa, 2017-18 (PHIRST): a population cohort study. Lancet Glob Health 2021; 9: e863–e874.

3 Olsen SJ, Winn AK, Budd AP, et al. Changes in Influenza and Other Respiratory Virus Activity During the COVID-19 Pandemic - United States, 2020-2021. MMWR Morb Mortal Wkly Rep 2021; 70: 1013–9.

4 Laurie KL, Rockman S. Which influenza viruses will emerge following the SARS-CoV-2 pandemic? Influenza Other Respir Viruses 2021; 15: 573–6.

5 Koutsakos M, Wheatley AK, Laurie K, Kent SJ, Rockman S. Influenza lineage extinction during the COVID-19 pandemic? Nat Rev Microbiol 2021; 19: 741–2.

6 Qi Y, Shaman J, Pei S. Quantifying the Impact of COVID-19 Nonpharmaceutical Interventions on Influenza Transmission in the United States. J Infect Dis 2021; 224: 1500–8.

7 Huang QS, Wood T, Jelley L, et al. Impact of the COVID-19 nonpharmaceutical interventions on influenza and other respiratory viral infections in New Zealand. Nat Commun 2021; 12: 1001.

8 Feng L, Zhang T, Wang Q, et al. Impact of COVID-19 outbreaks and interventions on influenza in China and the United States. Nat Commun 2021; 12: 3249.

9 Ali ST, Lau YC, Shan S, et al. Prediction of upcoming global infection burden of influenza seasons after relaxation of public health and social measures during the COVID-19 pandemic: a modelling study. Lancet Glob Health 2022; 10: e1612–22.

10 Dhanasekaran V, Sullivan S, Edwards KM, et al. Human seasonal influenza under COVID-19 and the potential consequences of influenza lineage elimination. Nat Commun 2022; 13: 1721.

11 Jones N. Why easing COVID restrictions could prompt a fierce flu rebound. Nature 2021; 598: 395.

12 Baker RE, Park SW, Yang W, Vecchi GA, Metcalf CJE, Grenfell BT. The impact of COVID-19 nonpharmaceutical interventions on the future dynamics of endemic infections. Proc Natl Acad Sci U S A 2020; 117: 30547–53.

13 World Health Organization (WHO). FluNet. Available at: https://www.who.int/tools/flunet.

14 World Health Organization (WHO). FluID. Available at: https://www.who.int/teams/global–influenza–program.

15 EuroMOMO Network.. EuroMOMO Winter Season 2015/16 Mortality Summary Report. Available at: https://www.euromomo.eu/uploads/pdf/winter_season_summary_2015_16.pdf.

16 Nielsen J, Vestergaard LS, Richter L, et al. European all-cause excess and influenza-attributable mortality in the 2017/18 season: should the burden of influenza B be reconsidered? Clinical Microbiology and Infection 2019; 25: 1266–76.

17 van Bilsen WPH, Boyd A, van der Loeff MFS, et al. Diverging trends in incidence of HIV versus other sexually transmitted infections in HIV-negative MSM in Amsterdam. AIDS 2020; 34: 301–9.

18 Wynberg E, van Willigen HDG, Dijkstra M, et al. Evolution of Coronavirus Disease 2019 (COVID-19) Symptoms During the First 12 Months After Illness Onset. Clinical Infectious Diseases 2021; Published online Sept. DOI:10.1093/cid/ciab759.

19 Lam EKS, Morris DH, Hurt AC, Barr IG, Russell CA. The impact of climate and antigenic evolution on seasonal influenza virus epidemics in Australia. Nat Commun 2020; 11: 2741.

20 Ohmit SE, Petrie JG, Cross RT, Johnson E, Monto AS. Influenza Hemagglutination-Inhibition Antibody Titer as a Correlate of Vaccine-Induced Protection. J Infect Dis 2011; 204: 1879–85.

21 Tsang TK, Perera RAPM, Fang VJ, et al. Reconstructing antibody dynamics to estimate the risk of influenza virus infection. Nat Commun 2022; 13: 1557.

22 Coudeville L, Bailleux F, Riche B, Megas F, Andre P, Ecochard R. Relationship between haemagglutination-inhibiting antibody titres and clinical protection against influenza: development and application of a bayesian random-effects model. BMC Med Res Methodol 2010; 10: 18.

23 Bedford T, Suchard MA, Lemey P, et al. Integrating influenza antigenic dynamics with molecular evolution. Elife 2014; 3: e01914.

24 Axelsen JB, Yaari R, Grenfell BT, Stone L. Multiannual forecasting of seasonal influenza dynamics reveals climatic and evolutionary drivers. Proceedings of the National Academy of Sciences 2014; 111: 9538.

25 Fonville JM, Wilks SH, James SL, et al. Antibody landscapes after influenza virus infection or vaccination. Science 2014; 346: 996–1000.

26 Belongia EA, Sundaram ME, McClure DL, Meece JK, Ferdinands J, VanWormer JJ. Waning vaccine protection against influenza A (H3N2) illness in children and older adults during a single season. Vaccine 2015; 33: 246–51.

27 Puig-Barberà J, Mira-Iglesias A, Tortajada-Girbés M, et al. Waning protection of influenza vaccination during four influenza seasons, 2011/2012 to 2014/2015. Vaccine 2017; 35: 5799–807.

28 Ranjeva S, Subramanian R, Fang VJ, et al. Age-specific differences in the dynamics of protective immunity to influenza. Nat Commun 2019; 10: 1660.

29 Department of Health and Aged Care. Australian Government. Australian Influenza Surveillance Report No. 14, 2022. Available at: https://www1.health.gov.au/internet/main/publishing.nsf/Content/B4F29E7D594818EBCA2588DB000EA9D2/$File/flu-14-2022.pdf (Accessed Oct 25, 2022).

