## Supplementary Material for "Potential impacts of prolonged absence of influenza virus circulation on subsequent epidemics"

### Supplementary Files


| | |
|---|---|
| **Supplementary Methods** | **2** |
| Epidemic composition data | 2 |
| Epidemic size data | 2 |
| Statistical modeling | 2 |
| Severity data | 3 |
| Viruses | 3 |
| Longitudinal serum samples | 4 |
| Haemagglutination inhibition assay | 4 |
| Antibody waning model | 5 |
| References | 6 |
| **Supplementary Figures** | **7** |

### Methods

#### Epidemic composition data

We downloaded records of virological surveillance data from the WHO FluNet[1] database for all countries in the temperate Northern and Southern Hemisphere from 2002 until 2020, or a shorter period for a limited subset of countries. We limited the dataset to countries in temperate zones because the discrete seasonal nature of flu epidemics in temperate zones facilitates the computation of lull durations and epidemic sizes. For each country, we retained the longest sequence of consecutive seasons in which at least 20 specimens were influenza-positive. In each season, defined as the period from the period from week 40 until week 20 for the Northern Hemisphere and the entire year for the Southern Hemisphere, we computed the proportion of all positive tests that was attributable to each of A/H3N2, pandemic A/H1N1pdm09 (from the 2009 pandemic onwards), and influenza B viruses. We did not break down influenza B viruses by lineage because in many countries influenza B viruses were not further characterized.

In many seasons, only a proportion of all influenza A virus positive tests were subtyped; in those cases, we approximated the total proportion of each subtype by assuming that the subtype of the non-subtyped influenza A virus specimens were distributed according to the relative proportions of subtyped influenza A viruses. Additionally, we required that in each country-season at least 20 positive specimens were characterized (though most counts were much higher). This resulted in a dataset of 718 season-country records over a period of 18 seasons, for 47 countries. We assigned a binary variable to each subtype/type in each season for dominance; we defined dominance as a subtype/type accounting for at least 30% of all detections in a country in a season. Hence, in principle, all three subtypes/types considered can be simultaneously dominant in a single season. To avoid including effects of the COVID-19 pandemic on influenza dynamics, we truncated the 2019-2020 season at the 15th of February 2020, and to avoid including the effect of the 2009 A/H1N1pdm09 pandemic, we truncated the 2008-2009 influenza season at the 1st of April 2009.

#### Epidemic size data

To estimate epidemic sizes for each subtype/type, we extracted weekly records of influenza-like illness from the WHO FluID[2] database. We limited this dataset to countries for which influenza-like illness (ILI) records were available for all seasons from 2010-2011 until 2019-2020, and for which virological surveillance data was available, as described above. Additionally, we required ILI curves to follow the expected shape of an influenza epidemic curve, i.e. peaking in winter and only sporadic isolation outside this period, and without periods of missing data. To facilitate the estimation of season effects, we only considered countries located in the Northern hemisphere. This yielded a set of 20 countries, each with 10 seasons worth of ILI data, located in Europe and the Middle East.

We approximated the relative epidemic size of each subtype in each country per season by multiplying total ILI incidence in that country's season by the proportion of all detections from that country in that season attributable to that (sub)type in the virological surveillance data, yielding a measure of (sub)type-specific ILI. We then computed the relative size of each (sub)type's epidemic in each country by computing the proportion of all ILI in that country in the total study period, i.e. from the 2010-2011 until 2019-2020 seasons, that was attributable to that (sub)type-specific ILI value, and multiplying this number by the total number of seasons. Hence, if in a particular country a (sub)type-specific epidemic had a relative size of 0.8, its size corresponded to 80% of the mean influenza epidemic size (irrespective of (sub)type) in that country in the ten-year period. In this way, this metric accounts for differences between seasons with regard to epidemic size, as opposed to only composition. We accounted for the COVID-19 pandemic as described above.

#### Statistical modeling

We used Bayesian hierarchical linear regression to estimate the effects of lull periods on epidemic size. As predictors, we separately used each of 1) years since previous dominance, 2) previous season size, or 3) previous two seasons' size sum. The first model has relative size as outcome and time since dominance as calculated using the virological surveillance data as predictor (N=188, 198, 180 for A/H3N2, A/H1N1pdm09 and B):

$$y_i \sim N(\mathrm{a} + \beta x_i, \sigma_y)$$

where $y_i$ is an epidemic's relative size for a certain (sub)type in country-season pair $i$, $a$ is the model intercept for that (sub)type, $\beta$ is the coefficient for number of years since dominance of the (sub)type, and $\sigma_y$ is the error standard deviation. $x_i$ represents the number of years since previous dominance of the (sub)type in country-season pair $i$ minus one, such that $a$ represents the predicted size if the previous dominance was in the previous year.We put weakly informative priors on the main effect, the intercept and the standard deviation.

$$\beta \sim N(0,1)$$

$$\mathrm{a} \sim N(0.5,1)$$

$$\sigma_y \sim Half - Normal(0,1)$$

We also ran the same models with season effects, where we furnished each season with its own intercept:

$$y_i \sim N(\mathrm{a}_{s[i]} + \beta x_i, \sigma_y)$$

Here, $a_{s[i]}$ is the season effect for that (sub)type corresponding to that season, for a country-season pair $i$. Season effects are shared between countries in a single season. We assumed that the season effects $\mathrm{a}_s$, constrained to positive values, are distributed according to a common mean $\mu_\mathrm{a}$ and common standard deviation $\sigma_\mathrm{a}$,and we put weakly informative priors on the mean season effect and its standard deviation:

$$\mathrm{a}_s \sim N(\mu_\mathrm{a}, \sigma_\mathrm{a})$$

$$\mu_\mathrm{a} \sim N(0.5,1)$$

$$\sigma_a \sim Half - Normal(0,1)$$

In addition to the model described above, we ran models with relative size as outcome and relative size in the previous year as the predictor (N=180 for each (sub)type) or the sum of the two previous years' sizes (N=160 for each (sub)type). We used the same model specification and priors as for the size~years since dominance model, but we replaced the predictor with the relative size in the previous season, or with the sum of relative size in the two previous seasons. All the above models were run both with and without season effects, *i.e.* with either a single value for the intercept, or with a separate intercept for each season. In the models with season effects, the season effects correspond to the predicted 'base size' of a (sub)type's epidemic in a particular season, given that either the previous dominance was in the previous year, there was no circulation in the previous year, or there was no circulation in the previous two years, respectively, for the models with years since previous dominance, previous season size, and previous two seasons' size sum as predictors. We ran the models for each (sub)type individually, and for each predictor individually. The models were fit using MCMC in Stan v2.21.0. The models were each run for 3000 iterations, discarding the first 1000 as burn-in, with four independent chains. All models were fit using MCMC in Stan v2.21.0, with convergence assessed by inspection of Rhat (< 1.05), effective sample size (> 200) and the trace plots. We compared models with and without season effects using leave-one-out cross-validation.[3]

**Severity data**

We used excess deaths as a proxy for epidemic severity. We extracted pooled Europe-wide winter-period excess mortality for the 2010-2011 and 2011-2012 seasons, and pooled-Europe wide flu-specific winter-period excess mortality for the 2012-2012 to 2017-2018 seasons from the EuroMOMO network, which combines excess deaths in European countries to estimate Europe-wide excess mortality.[4,5] The number of countries included in the calculation of excess deaths ranged from 7 in 2010/2011 to 24 in 2017/2018. For seasons that were 1) dominated by a single (sub)type in most countries; and 2) were uniform across most countries with regard to the number of years since previous dominance of the dominant (sub)type, we compared the Europe-wide excess mortality to the value for years since previous dominance. Because these criteria were only met for a small number of seasons, the comparison was performed mostly qualitatively.

**Viruses**

To select the four representative strains used in this study (A/Netherlands/04189/2017 (A/H3N2), A/Netherlands/10218/2018 (A/H1N1pdm09), B/Netherlands/04136/2017 (B/Yamagata), and

B/Netherlands/00302/2018 (B/Victoria)), we downloaded high-quality (<5% ambiguous nucleotides, >95% full length) seasonal influenza virus haemagglutinin sequences (A/H3N2, N=1,396; A/H1N1pdm09, N=1,283; B/Yamagata, N=1,129; and B/Victoria, N=1,408) collected between 2016 and October 2021 from GISAID (www.gisaid.org) and reconstructed maximum-likelihood phylogenetic trees for each influenza virus subtype using the general time reversible substitution model with IQ-TREE.[6] These trees were used to assess the representativeness of viruses from the Netherlands in the early portion of the study period and the selected viruses were all representative of viruses that caused epidemics in the Netherlands during the 2017/18 winter.

All four viruses were propagated in Madin-Darby Canine Kidney (MDCK) cells in infection medium which consisted of MEM-Eagle Medium /EBSS (Lonza, Geleen, The Netherlands) supplemented with MEM Non-Essential Amino Acids (Gibco, ThermoFischer Scientific, Amsterdam, The Netherlands), penicillin (100 U/mL), streptomycin (100 g/mL), L-Glutamine (Lonza), HEPES (Lonza), and TPCKtrypsin (Sigma-Aldrich/Merck, Darmstadt, Germany). They were harvested after 72 hours of incubation at either 37°C (H3N2 and H1N1) or 33°C (Yamagata and Victoria) and checked by Sanger sequencing.

**Longitudinal serum samples**

A total of 630 serum samples from 165 healthy male and female adults, including people >70 years of age (elderly), were collected in the Netherlands, longitudinally, before and during the COVID-19 pandemic in two separate cohorts: 1. Amsterdam Cohort Studies on HIV infection and AIDS[7](ACS) and 2. the Viro-immunological, clinical and psychosocial correlates of disease severity and long-term outcomes of infection in SARS-CoV-2 – a prospective cohort study[8]. The initial aim of the Amsterdam Cohort Studies was to investigate the prevalence, incidence, and risk factors of HIV-1 infection. The study population consists of men who have sex with men and live mainly around the city of Amsterdam, the Netherlands. Participation in ACS is voluntary and without incentive. Written informed consent of each participant was obtained at enrolment. The Amsterdam Cohort Studies on HIV infection and AIDS was approved by the Medical Ethics Committee of the Amsterdam University Medical Centre of the University of Amsterdam, the Netherlands (MEC 07/182). Participants from the ACS cohort included in our study were all HIV-1 seronegative men ranging from 22 to 70 years old at the time of sample collection in mid-2017. Briefly, five stored samples were used per participant, i.e. 1. mid-2017, 2. mid-2018, 3. mid-2019, 4. mid-2020, 5. mid-2021.

Data derived from the ACS participants was complemented by the data acquired from participants in the SARS-CoV-2 cohort RECoVERED. The aim of the RECoVERED cohort study is to describe the immunological, clinical and psychosocial sequelae of a SARS-CoV-2 infection. Individuals aged 16 to 85 years with laboratory-confirmed SARS-CoV-2 infection were enrolled on May 2020 till the end of June 2021 in the municipal region of Amsterdam, the Netherlands. All participants provided written informed consent. The RECoVERED study was approved by the medical ethical review board of the Amsterdam University Medical Centre (NL73759.018.20). From RECoVERED, we selected a total of 65 individuals, both male and female adults ranging from 20 to 77 years old at the time of sample collection in mid-2020, all of which had a confirmed SARS-CoV-2 infection but were otherwise healthy and unvaccinated for influenza in 2020. For these 65 individuals, samples were collected in the summer period of 2020 and 2021 only (two total for each participant).

**Hemagglutination inhibition (HI) assay**

All serum samples were receptor destroying enzyme (RDE)-treated, as described elsewhere.[9] Briefly, 100mL of serum samples from ACS individuals 1-30 were combined with 200mL of RDE; for serum samples from ACS individuals 31-100 and all 65 RECoVERED subjects, 100mL of serum were combined with 300mL of RDE. This difference in protocol was per the instructions of the providers of the respective batches of RDE. Because of this protocol difference, the results of ACS participants 1-30 and 31-100 are shown separately. All samples were then incubated at 37°C for 18 to 20 hours. The RDE reaction was then halted by heating the treated samples at 56°C for 30 to 60 minutes.

The hemagglutination inhibition activity of all serum samples was tested in an hemagglutination inhibition assay as described elsewhere[9] using two replicates per sample for A/H1N1pdm09, B/Yamagata, and B/Victoria viruses, and one single measurement for A/H3N2 viruses. Due to inefficient agglutination of turkey red blood cells (tRBCs) by recent A/H3N2 viruses, we used glycan remodeled tRBCs expressing appropriate receptors for recent A/H3N2 virusesfor the HI assays of the A/H3N2 virus stock.[10] Briefly, the hemagglutination titer of each of the four viruses was determined by doing a two-fold serial dilution of 50mL of each virus stock and adding 50mL of PBS and 25mL of 1% turkey red blood cells (tRBCs) to each well, followed by one hour incubation at 4°C and the reading of the hemagglutination patterns. The virus stocks were then diluted to a concentration of four hemagglutination units (HAU). The diluted viruses were then incubated with 50mL of two-fold serially diluted serum, in a total volume of 75mL for 30 minutes at 37°C. The initial dilution used for the serial dilution of the serum was 1:20 of the RDE treated serum. After the incubation step, 25mL of 1% turkey red blood cells were added to the serum-virus mix and incubated at 4°C for one hour. The hemagglutination inhibition patterns were then read out and used for the calculation of antibody titers.

**Antibody waning model**

For the RECoVERED cohort for the years 2020 and 2021, all participants were confirmed to have not received an influenza vaccination between the two sample collections and no natural influenza infection can be safely assumed given the near absence of influenza in the Netherlands during this period. For the ACS data, those who experienced a four or greater fold increase in titer between consecutive visits for a particular strain had their strain-data discarded in order to remove the obscuring effects of vaccination and infection. Due to an apparent batch effect, the ACS individuals were split into two separate groups of 70 and 30 individuals. Results from the larger group are presented in the main text while the smaller group of 30 is referred to the Supplementary Material (Supplementary Fig. 4).

True antibody titer $\log_2$ HI, $\tilde{T}^i$, as opposed to that measured by hemagglutination inhibition assay, $T^i$, is a continuous variable which we assume, for every individual, $i$, decays with time, $t$, as

$$\tilde{T}^i = c^i - \alpha^i t$$

Where $c^i$ are individual specific initial titers and $\alpha^i$ are the individual waning rates. The waning rates are assumed to be normally distributed about a population mean, $\alpha_\mu$, with standard deviation, $\alpha_\sigma$.

If serum dilutions could be performed in arbitrarily small increments, we assume the point at which hemagglutination would be observed to cease, $T_{obs}$, to be distributed normally about the true value:

$$T_{obs} \sim N(\tilde{T}, \epsilon)$$

where $\epsilon$ shall be referred to as the "measurement error". Instead, with discrete dilutions in increments of one, the probability of measuring $T \in \{0, 1, 2...8\}$ is the probability that $T_{obs}$ falls between $T$ and $T-1$. Thus, the measurement probability is given by:

$$P(T \mid \tilde{T}, \epsilon) = \begin{cases} \Phi(1, \tilde{T}, \epsilon) & T < 1 \\ \Phi(\mathrm{T}, \tilde{T}, \epsilon) - \Phi(T_m - 1, \tilde{T}, \epsilon) & 1 \leq T < 8 \\ 1 - \Phi(8, \tilde{T}, \epsilon) & T \geq 8 \end{cases}$$

where $\Phi(x, \mu, \sigma)$ is the cumulative distribution function of the normal distribution.

Our data for each individual, $i$, consists of a series of titer measurements, $\boldsymbol{T}^{i,r} = (T_1^{i,r}, T_2^{i,r}, ...., T_n^{i,r})$, at corresponding timepoints $\boldsymbol{t}^i = (t_1^i, t_2^i, ...., t_n^i)$, where $r \in \{1,2\}$ indicates replicate measurements. To infer the probability of the unknown parameters $\epsilon$, $\alpha_\mu$ and $\alpha_\sigma$ given the data, it is necessary to augment the data by introducing individual intercepts. For one replicate from one individual, the likelihood of unknown parameters $\alpha_\mu$, $\alpha_\sigma$, $\epsilon$, and $c^i$ then becomes:

$$p(\alpha_\mu, \alpha_\sigma, \epsilon, c^i | T^{i,r}) \propto p(T^{i,r} |, \alpha_\mu, \alpha_\sigma, \epsilon, c^i) \Pi(, \alpha_\mu, \alpha_\sigma, \epsilon, c^i)$$

$$\propto p(T^{i,r} | \tilde{\boldsymbol{T}}^i (\alpha_\mu, \alpha_\sigma, \epsilon, c^i)) \Pi(\alpha_\mu, \alpha_\sigma, \epsilon, c^i)$$

where $\tilde{\boldsymbol{T}}^i(\alpha_\mu, \alpha_\sigma, \epsilon, c^i) = (\tilde{T}_1^{i,r}, \tilde{T}_2^{i,r}, ...., \tilde{T}_n^{i,r})$ are the true values of titer, given the unknown parameters and $\Pi$ is the prior joint distribution of the parameters. The total log likelihood is thus the sum over all individuals and replicates:

$$L(\alpha_\mu, \alpha_\sigma, \epsilon, c | \boldsymbol{T}) \propto \sum_i \sum_r log(p(\alpha_\mu, \alpha_\sigma, \epsilon, c^i | \boldsymbol{T}^{i,r}))$$

A Markov Chain Monte Carlo (MCMC) algorithm implemented in Stan v2.21.0 was used to explore the distribution of model parameters (waning and measurement) and augmented data (individual intercepts). This model was run on four independent chains, each consisting of 5,000 iterations discarding the first 2,500 as burn in. Weakly-informative priors were used and convergence was assessed by inspection of the trace plots and Rhat. Analyses were conducted using R v4.0.3, with code available in the GitHub repository.

References


1 World Health Organization (WHO). FluNet. Available at: https://www.who.int/tools/flunet.

2 World Health Organization (WHO). FluID. Available at: https://www.who.int/teams/global–influenza–program.

3 Vehtari A, Gelman A, Gabry J. Practical Bayesian model evaluation using leave-one-out cross-validation and WAIC. *Stat Comput* 2017; **27**: 1413–32.

4 EuroMOMO Network. EUROMOMO WINTER SEASON 2015/16 MORTALITY SUMMARY REPORT. https://www.euromomo.eu/uploads/pdf/winter_season_summary_2015_16.pdf.

5 Nielsen J, Vestergaard LS, Richter L, *et al.* European all-cause excess and influenza-attributable mortality in the 2017/18 season: should the burden of influenza B be reconsidered? *Clinical Microbiology and Infection* 2019; **25**: 1266–76.

6 Minh BQ, Schmidt HA, Chernomor O, *et al.* IQ-TREE 2: New Models and Efficient Methods for Phylogenetic Inference in the Genomic Era. *Mol Biol Evol* 2020; **37**: 1530–4.

7 van Bilsen WPH, Boyd A, van der Loeff MFS, *et al.* Diverging trends in incidence of HIV versus other sexually transmitted infections in HIV-negative MSM in Amsterdam. *AIDS* 2020; **34**: 301–9.

8 Wynberg E, van Willigen HDG, Dijkstra M, *et al.* Evolution of Coronavirus Disease 2019 (COVID-19) Symptoms During the First 12 Months After Illness Onset. *Clinical Infectious Diseases* 2021; published online Sept. DOI:10.1093/cid/ciab759.

9 WHO Global Influenza Surveillance Network Manual for the laboratory diagnosis and virological surveillance of influenza. 2011 https://apps.who.int/iris/bitstream/handle/10665/44518/9789241548090_eng.pdf?sequence=1.

10 Broszeit F, van Beek RJ, Unione L, *et al.* Glycan remodeled erythrocytes facilitate antigenic characterization of recent A/H3N2 influenza viruses. *Nat Commun* 2021; **12**: 5449.

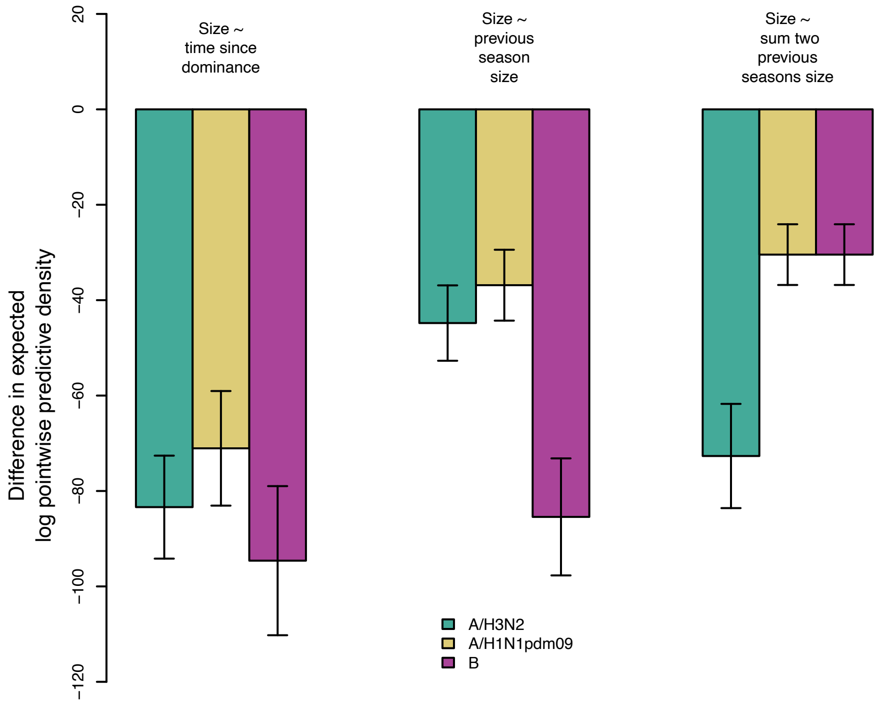


**Supplementary Fig. 1. Model comparison.** The difference in expected log posterior predictive density between the same models with and without season effects, for all three main model formulations discussed in the main text. Error bars indicate the standard error of the difference.

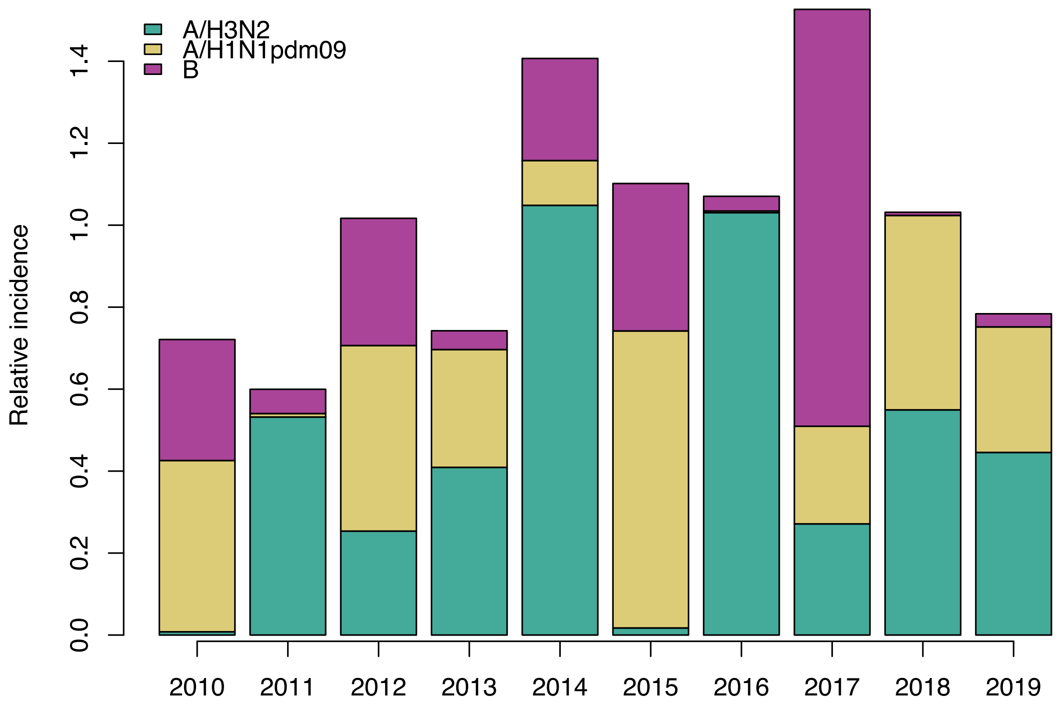


**Supplementary Fig. 2. Relative sizes of seasonal influenza epidemics in the Netherlands in the decade preceding the COVID-19 pandemic.** The relative size of influenza A/H3N2, A/H1N1pdm09, and B epidemics in the Netherlands, defined as the influenza-like illness rate in the Netherlands per season multiplied by the proportion of isolates corresponding to each (sub)type in each season, scaled such that a relative size of one corresponds to the mean total influenza-like illness rate in a single season.

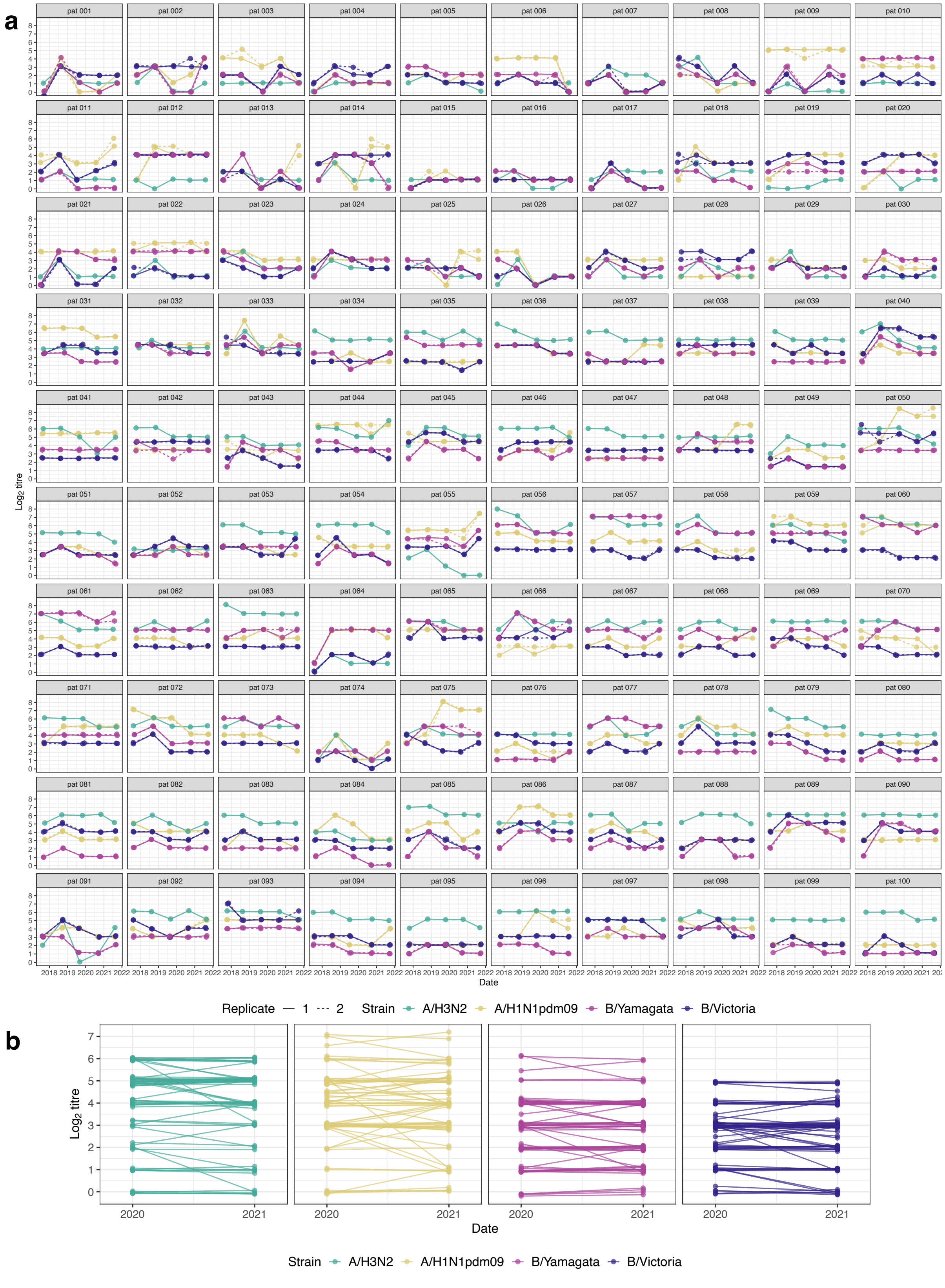


**Supplementary Fig. 3. Individual HI titre dynamics for two independent cohorts.**
**a,** Each panel depicts $\log_2$ individual HI titres from 2017-2021 for all participants in the Amsterdam Cohort Studies on HIV infection and AIDS (ACS). Strains are differentiated by colour and, where applicable, replicate measurements are indicated with dashed lines. **b,** Subtype-specific titre patterns, averaged over two replicates,

for all individuals within the RECoVERED cohort. Slight random jitter has been added to both panels in the x and y dimensions to avoid obscuring of repeated data points.

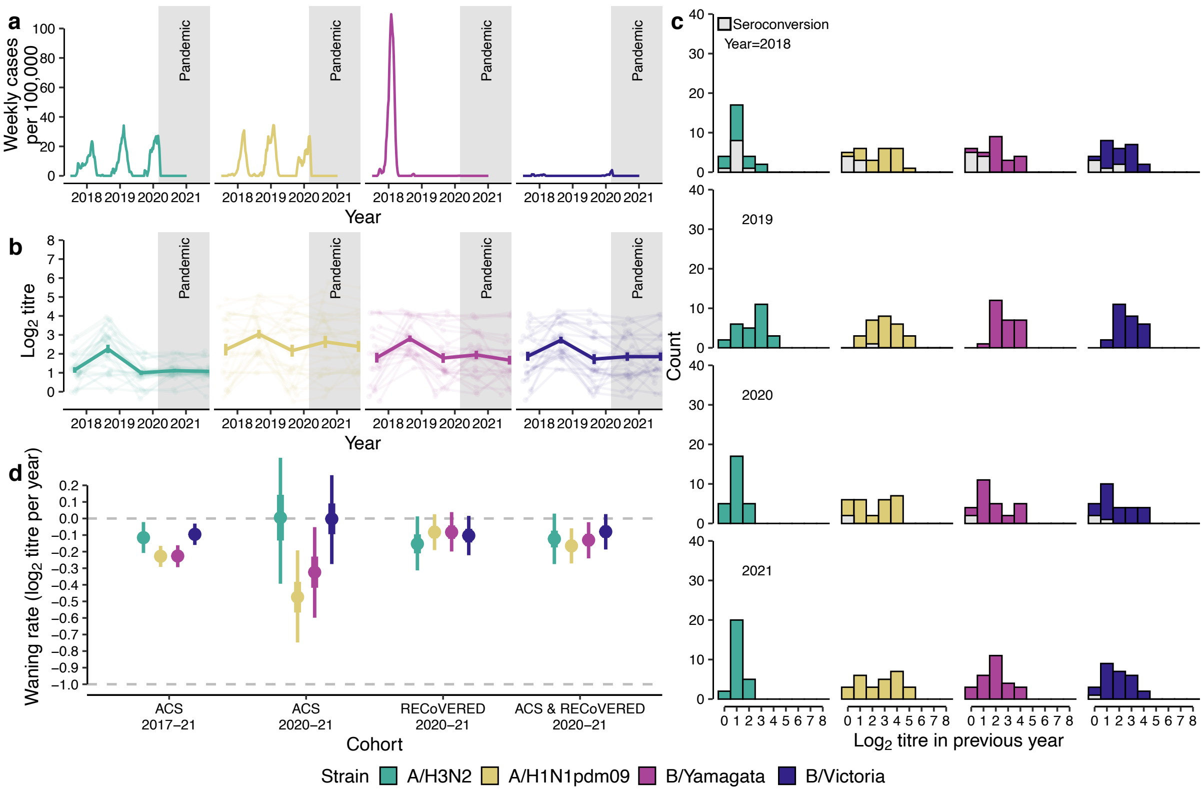


**Supplementary Fig. 4. Waning antibody titres to seasonal influenza virus before and during the COVID-19 pandemic for a subset of the Amsterdam Cohort Studies on HIV infection and AIDS (ACS).** The information content of this figure corresponds to that contained in Fig. 1, but for individuals 1-30 of the ACS cohort (see Methods. **a**, Seasonal influenza epidemic activity 2017-2021 in the Netherlands based on virological and syndromic surveillance data. **b**, Individual antibody titres against seasonal influenza viruses based on haemagglutination inhibition (HI) assay from 2017-2021 among 30 healthy male adult participants of the ACS cohort for each influenza virus (sub)type. Mean antibody tires changes across all individuals are drawn in bold lines with error bars indicating the mean standard error (n=30). **c**, HI titre distributions in the ACS cohort following each winter epidemic period coloured by influenza virus (sub)type. HI titre distributions of individuals who experienced a ≥2 log2 units increase in HI titre (≥4-fold increase in HI titre), indicating likely infection in the previous winter epidemic period, are shown in grey bars. **d**, Antibody waning rates by influenza virus type and subtype in adults estimated from HI titres from 30 ACS and 65 RECoVERED individuals. Error bars correspond to the 50% and 95% credible interval from the Markov Chain Monte Carlo algorithm used to explore the distribution of model parameters. Waning rate of -1.0 corresponds to one two-fold decrease in HI antibody titre in one year. Details on waning and measurement parameter estimates detailed in Extended Data Table. 2.
